# Qualitative assessment of SARS-CoV-2-specific antibody avidity by lateral flow immunochromatographic IgG/IgM antibody assay

**DOI:** 10.1101/2020.06.23.20138016

**Authors:** Arantxa Valdivia, Ignacio Torres, Dixie Huntley, María Jesús Alcaraz, Eliseo Albert, Javier Colomina, Josep Ferrer, Arturo Carratalá, David Navarro

**Affiliations:** Microbiology Service, Hospital Clínico Universitario, INCLIVA Research Institute, Valencia, Spain; Medical Biochemistry and Clinical Analysis Service, Hospital Clínico Universitario, INCLIVA Research Institute, Valencia, Spain; Department of Microbiology, School of Medicine, University of Valencia, Valencia, Spain

**Keywords:** SARS-CoV-2, Covid-19, antibodies, avidity, urea dissociation

## Abstract

Qualitative assessment of SARS-CoV-2-specific antibody avidity was conducted using an urea (6M) dissociation test performed on a lateral flow immunochromatographic IgG/IgM device. We included a total of 76 serum specimens collected from 57 COVID-19 patients, of which 39 tested positive for both IgG and IgM and 37 only for IgG. Sera losing IgG reactivity after urea treatment (n=28) were drawn significantly earlier (*P*=0.04) after onset of symptoms than those which preserved it (n=48). This assay may be helpful to estimate the time of acquisition of infection in patients with mild to severe COVID-19.

---

Detection of SARS-CoV-2 RNA in respiratory tract specimens by reverse-transcription-based PCR (RT-PCR) assays is the mainstay of COVID-19 diagnosis [1]. However, a non-negligible fraction of COVID-19 patients test negative by RT-PCR on initial or consecutive upper respiratory tract specimens, due to a number of non-mutually exclusive pre-analytical or analytical factors [2]. Although serology testing is mainly aimed at identifying individuals who have previously been exposed to SARS-CoV-2, it may also aid in diagnosis of ongoing COVID-19, particularly in RT-PCR negative patients who present at relatively late times after infection [3]. Knowledge of the precise timing of infection may be of clinical and epidemiological relevance as viral shedding in the upper respiratory tract (URT) seems to continue up to 7-9 days after onset of symptoms in patients presenting with mild or moderate COVID-19 [4-6]. Often enough, however, this cannot be accurately determined. Theoretically, virus-specific serum IgM antibodies appear as soon as 7 days after infection and precede IgG seroconversion [7]. Nevertheless, both synchronous seroconversion of IgG and IgM, and IgM seroconversion occurring later than IgG have been documented in the setting of COVID-19 [8], casting doubt on the reliability of SARS-CoV-2 IgM as a biomarker of acute infection. Affinity maturation is a process by which Th_2_-cell-activated B cells produce IgG antibodies with increased affinity for the antigen during the course of an immune response [9], and avidity is defined as the combined affinities of a mixture of polyclonal IgG molecules [10]. Presence of low-avidity IgGs has conventionally been considered an indicator of recent infection [10]. Here, we carried out qualitative assessment of SARS-CoV-2-specific antibody avidity using an urea dissociation test performed on a lateral flow immunochromatographic device-LFIC-[11], and also discuss the potential clinical use of this approach.

A total of 76 serum specimens collected from 57 COVID-19 patients were included in this study. Microbiological diagnosis of COVID-19 was made by RT-PCR in 44 patients, using commercially-available RT-PCRs on upper respiratory tract specimens (URT) [12,13], and by lateral flow immunochromatographic (LFIC) assay in the remaining 13 patients (described below). Median age of patients (32 male and 25 female) was 66 years (range, 27-99 years). Forty-seven patients were admitted to our center with pneumonia, while the remaining 10 patients presented with mild symptoms not requiring hospitalization. Comorbid conditions including diabetes, cardiovascular diseases, chronic obstructive pulmonary disease or malignancies were identified in 47 patients. Clinical charts were reviewed to establish the time of onset of symptoms. The current study was approved by the Ethics Committee of Hospital Clínico Universitario INCLIVA.

Qualitative assessment of SARS-CoV-2 antibody avidity was carried out using the ALLTEST 2019-nCoV IgG/IgM Rapid Test Cassette (Hangzhou ALLTEST Biotech Co., Ltd. Hangzhou, China), which uses a recombinant SARS-CoV-2 N protein as the antigen, following the manufacturer’s recommendations [14]. Cryopreserved specimens (−80°C) were thawed and used for the experiments. A volume of 10µL of serum was diluted into 1 mL of sample buffer before depositing (100 µL) into the appropriate location of the cassette (Test T-hole). When the fluid was about to reach the absorbent pad, 100 µL of sample buffer containing 6M urea was added to the T hole on the card. Serum specimens were run in parallel in the absence of urea treatment. Each reading was carried out independently by two observers after 20 min incubation. Appearance of either strong or weak sharp bands at the T line was recorded as a positive result. Absence of discernible lines was recorded as negative. Complete disappearance of reactive lines after urea treatment was interpreted as presence of low-avidity antibodies, whereas their persistence was taken to indicate high-avidity antibody presence.

The Mann-Whitney U-test was used for comparison of medians. *P* values <0.05 were deemed to be statistically significant. The statistical package SPSS, version 21.0 (SPSS Inc., USA) was employed.

Out of the 76 sera, 39 tested positive for both IgG and IgM and 37 only for IgG. IgG+/IgM+ and IgG+/IgM-sera were obtained at a median of 20 days (range, 1-48 days) and 24 days (range, 6-59) after onset of symptoms, respectively (*P*=0.13). In line with previous observations [8], our data highlighted the wide variability in the kinetics of IgM and IgG detection across Covid-19 patients, which detracted from the reliability of IgM presence as a marker of acute infection.

Following urea treatment, IgG reactivity disappeared in 28 sera and persisted in 48. Sera losing IgG reactivity were obtained significantly earlier (*P*=0.04) after onset of symptoms than those preserving it (median, 14.5 days; range, 1-45 days vs. median, 23 days; range, 5-59 days, respectively), although a certain degree of overlap was seen. Based upon the assumption that viable SARS-CoV-2 can be detected in URT specimens up to 9 days after symptoms onset [4-6], we grouped sera into two categories according to whether they were drawn either prior to (n=10) or after day 9 (n=66) since symptoms onset. Low-avidity IgGs were detected in 8 out of the 10 former sera, and in 21 of the 66 latter sera (*P*=0.01).

At least two consecutive sera were available from 15 patients (Table 1). Acquisition of high-avidity IgG antibodies in our system was clearly time-dependent, usually occurring 3 weeks after onset of symptoms, although in a few patients it could be documented earlier (patients 4, 5 and 15). This observation is in keeping with the idea that the dynamics of antibody affinity maturation varies across individuals [7,10].

IgM reactivity was lost in 17 out of 39 sera after urea dissociation treatment, whereas it remained in 22. The time elapsed since onset of symptoms did not differ across comparison groups (median 24 days; range 1-45 days vs. 14.5 days; range 2-48 days, respectively; *P*=0.14). Sera testing positive for rheumatoid factor (RF) IgM have been shown to yield false-positive IgM reactivity in a SARS-CoV-2 LFIC assay, and it has been reported that RF interference could be eliminated in most sera after urea dissociation [11]. This observation was validated in the current study: 5 out of 14 sera with leftover sample available that had lost IgM reactivity in the presence of urea tested positive for RF (median 18 IU/ml; range, 17 to 46 IU/ml; normal values <14 IU/ml). Collectively, these data suggest that SARS-CoV-2 N protein-reactive IgM avidity may vary across Covid-19 patients, regardless of the infection acquisition time; elucidation of whether this variability may impact on IgM functionality, and ultimately on Covid-19 prognosis could be an interesting focus of future research.

A total of 9 sera from patients with seasonal human coronavirus infection occurring prior to the epidemic outbreak in our Health Department were also included in the current study. Coronaviruses were detected in URT specimens by a multiplex PCR assay (The NxTAG® Respiratory Pathogen Panel; Luminex Corp, Austin, TX, USA). Seven patients had coronavirus 229E and 2 patients had dual infections caused by coronavirus 229E and HKU1 and coronavirus 229E and NL63. Sera had been obtained at a median of 3 weeks after diagnosis. Only one of the 9 sera tested IgG-positive, yet reactivity was lost following urea treatment. This suggested, but did not prove, that IgG antibodies targeting seasonal coronaviruses and cross-reacting with SARS-CoV-2 may display low avidity.

In summary, we adapted a commercially-available LFIC IgG/IgM assay for qualitative assessment of SARS-CoV-2 antibody avidity that may be helpful to estimate the time of acquisition of infection in patients with mild to severe COVID-19. Our approach should be validated using conventional quantitative ELISA or CLIA avidity assays and also in cohorts including both asymptomatic and paucisymptomatic individuals. Further studies are warranted to elucidate how the kinetics of IgG avidity maturation correlates with that of virus excretion in the URT, to determine the extent to which contagiousness of COVID-19 patients can be inferred from absence of high-avidity IgGs and determine whether avidity of SARS-CoV-2-specific IgGs impact on clinical outcomes.

## Data Availability

The data that support the findings of this study are available from the corresponding author, [author initials], upon reasonable request

## Acknowledgements

We are grateful to all personnel working at Microbiology Service of Hospital Clínico Universitario for their unwavering commitment in the fight against Covid-19. We are indebted to all colleagues who attended the patients, in particular to Josep Redón, María Luisa Blasco, Jaime Signes-Costa, María José Galindo, and María José Forner.

## Compliance with ethical standards

### Ethical approval

The current study was approved by the Ethics Committee of Hospital Clínico Universitario INCLIVA (2020-04).

### Informed consent

Written informed consent was waived by the Ethics committee.

